# Predicting Depression from Hearing Loss Using Machine Learning

**DOI:** 10.1101/2020.08.31.20185421

**Authors:** Matthew G. Crowson, Kevin H Franck, Laura C. Rosella, Timothy C. Y. Chan

## Abstract

**Background:** Hearing loss is the most common sensory loss in humans and carries an enhanced risk of depression. No prior studies have attempted a contemporary machine learning approach to predict depression using subjective and objective hearing loss predictors.

**Objective:** To deploy supervised machine learning to predict scores on a validated depression scale using subjective and objective audiometric variables and other health determinant predictors.

**Design:** We used a large predictor set of health determinants to forecast individuals’ scores on a validated instrument to screen for the presence and severity of depression (PHQ-9). After model training, the relative influence of individual predictors on depression scores was stratified and analyzed.

**Setting:** National Health and Nutrition Examination Survey (NHANES) 2015-2016 database.

**Patients:** Adult survey participants.

**Measurements:** Model prediction error performance.

**Results:** The test-set mean absolute error was 3.03 (CI 95%: 2.91-3.14) and 2.55 (CI 95%: 2.48-2.62) on datasets with audiology-only predictors and all predictors, respectively, on the PHQ-9’s 27-point scale. Participants’ self-reported frustration when talking to members of family or friends due to hearing loss was the fifth-most influential of all predictors. Of the top ten most influential audiometric predictors, five were related to social contexts, two for significant noise exposure, two objective audiometric parameters, and one presence of bothersome tinnitus.

**Conclusions:** Machine learning algorithms can accurately predict PHQ-9 depression scale scores from NHANES data. The most influential audiometric predictors of higher scores on a validated depression scale were social dynamics of hearing loss and not objective Such models could be useful in predicting depression scale scores at the point-of-care in conjunction with a standard audiologic assessment.

## INTRODUCTION

Hearing loss is the number one sensory loss in humans.(1) The functional associations of hearing loss include detriments to quality of life(2,3), cognition(4–7), social isolation(8–10), and mental health(11–24). Within the domain of mental health, a significant risk of depression in individuals with hearing loss has been demonstrated in several cohorts composed of adults spanning all ages and both sexes.(11,16,19–21,23) The severity of hearing loss has also been shown to be positively associated with depression severity.(18) Despite the link between hearing loss and depression, hearing aids have not been consistently shown to be effective in mitigating the effects of depression.(25–28) The failure to observe prevention or improvement in depression by treating hearing loss may be, at some level, due to a complex relationship between hearing loss and depression that we have yet to fully understand.

The National Health and Nutrition Examination Survey (NHANES) is a cross-sectional public health survey designed to assess the health and nutrition status of U.S. persons. NHANES data is advantageous for investigations seeking to analyze the interplay between determinants of health and specific medical conditions such as hearing loss due to the wide demographic representation of the subjects. In the past decade, several reports utilizing NHANES data reported on the prevalence of hearing loss in children and adults over time.(29–32) Other inferential studies revealed significant associations between hearing loss and risk of hospitalization in older adults(33), secondhand smoke(34), self-reported falls(35), and exposure to hazardous levels of occupational noise(36). Mener et al. demonstrated that hearing aid use was protective against a diagnosis of major depressive disorder using NHANES data, but hearing loss itself was not associated with a greater odds of major depressive disorder or depressive symptoms.(37) However, the study only included speech-frequency pure-tone average of hearing thresholds as a proxy measure for individuals’ total hearing loss experience. Moreover, no prior NHANES-based studies have attempted a predictive approach using subjective and objective hearing loss predictors.

Our primary hypothesis is that a predictive approach using machine learning and audiometric variables will perform with a high degree of accuracy and may be useful for identifying depression in patients with audiometric data. In the present study, we deploy interpretable supervised machine learning using NHANES data, composed of subjective and objective audiometric variables and several other health determinant predictors, to predict scores on a validated depression scale.

## METHODS

### NHANES Database Predictor Variables

The NHANES 2015-2016(38) dataset was mined for demographic (NHANES DEMO_I dataset), pre-existing medical conditions (MCQ_I), income (INQ_I), disability (DLQ_I), health insurance (HIQ_I), health care utilization and access (HUQ_I), occupation (OCQ_I), and physical functioning (PFQ_I) variables. Objective audiometry (AUX_I) testing results were also included, comprising otoscopy findings (normal – yes/no), excessive cerumen (yes/no), tympanometric data (width, volume, compliance, pressure), tympanogram type (A, AD, AS, B, or C), and bilateral pure tone thresholds (dB; hearing level) at 500 Hz, 1000 Hz, 2000 Hz, 4000 Hz, 6000 Hz, and 8000 Hz. We calculated the pure-tone average (PTA) for the left and right ears using the average threshold values of 500Hz, 1000Hz, and 2000Hz frequency datapoints. These PTAs were added to the dataset as new features. Functional audiometric content (AUQ_I) variables included self-reported hearing condition, satisfaction with hearing in social contexts, hearing health history, use of hearing assistive and protective devices, tinnitus history and exposure to occupational noise.

### Validated Depression Score Target Variable

The depression screener variables (DPQ_I) were composed of the validated Patient Health Questionnaire (PHQ-9) survey question responses.(39,40) Individual depression screening question scores were totaled to generate an overall PHQ-9 score. The total PHQ-9 score was used as our target variable. The total scores are interpreted as depression severity categories of ‘None’ (total score 1-4), ‘Mild’ (total score 5-9), ‘Moderate’ (total score 10-14), ‘Moderately Severe’ (total score 15-19), and ‘Severe’ (total score 20-27).(39,40)

### Modeling Approach

We trained an interpretable machine learning algorithm for a regression task to predict individuals’ PHQ-9 total score. We used the LightGBM algorithm, a variant of a gradient boosting decision tree architecture developed to enhance computational efficiency with high dimensional data.(41) A tree-based algorithm was chosen so we could interpret the model’s most influential features in predicting PHQ-9 scores. We ran two regression experiments. The first experiment included all NHANES predictor variables to predict a PHQ-9 total score. The second experiment included only the objective audiometric and subjective hearing loss NHANES predictors to predict the PHQ-9 total score.

### Data Preparation

The dataset used for the experiments was anchored to NHANES survey participants who had complete audiology predictor variables. Prior to analysis, the predictor variable content was screened to remove individual variables that might directly imply depression. For example, *DLQ_I variable #170* asked participants if they had “taking medication for depression.” We also removed all individual questions that comprise the PHQ-9 questionnaire. After removing these variables, the final predictor set comprised 151 numeric and categorical variables.

The data were pre-processed to impute missing numeric values using the variables’ mean, and drop variables with high multicollinearity (> 0.90 threshold). Power transforms were used to make data more normal-like: The predictors were transformed using the Yeo-Johnson method(42) and the PHQ-9 score target variable was transformed using the Box-Cox method.(43) Categorical variables were one-hot encoded, a transformation that facilitates inclusion of this data type into machine-learning algorithms.

### Model Training & Testing

A nested cross-validation approach was used for model training and testing. Cross-validation trains a new model on a subset of data, and validates the trained model on the remaining data. The inner loop employed 10-fold cross-validation for hyperparameter tuning on the training data, using 50 iterations within a nested random grid search.(44) For every grid iteration, the model randomly selected one value from the pre-defined grid of hyperparameters. The outer loop employed five-fold cross-validation on all data to evaluate the performance of the optimized models. Performance on validation and testing sets was measured using mean absolute error (MAE). The MAE in our experiment was the difference between the known PHQ-9 values and the models’ predicted PHQ-9 values. This process was repeated for the second experiment, which only used objective and subjective audiology predictors.

### Model Interpretation

To examine which predictors carried the highest influence in predicting total PHQ-9 score, the SHAP (Shapley Additive eXplanations) module was used quantify individual predictor importance.(45) SHAP assigns each predictor an importance value for a target prediction of a model. By assigning an importance value to each predictor, the SHAP provides an interpretable stratification of individual predictor influence relative to other predictor variables. (45)

### Hardware & Software

Data processing and the machine learning analyses were completed using R (R Core Team (2013), Vienna, Austria), and the PyCaret python package (1.0.0, pycaret.org). The model training and experiments were completed on a custom-built desktop computer with an AMD Ryzen Threadripper 1950X processor, 64GB of read-only memory (RAM), and a GeForce RTX 2070 8GB graphics card.

## RESULTS

### Descriptive Statistics

After joining the audiology predictors with other NHANES predictors of interest, the experimental cohort comprised 4,582 respondents. The mean age of the respondents was 44.2 years (std dev. 14.3), and 52.5% were female (n = 2,406). The mean PHQ-9 total score was 5.00 (std dev 4.80, range 1-27), with 1,755 (61.5%) of participants having ‘None,’ 696 (24.4%) ‘Mild’ depression severity, 237 (8.3%) moderate depression severity, 88 (3.1%) moderately-severe, and 74 (2.8%) severe (**Table 1**). The remaining descriptive statistics for all other predictors are available in the **Online Supplemental Results File**.

**Table 1.** Target variable descriptive statistics in 2015-2016 NHANES respondents who had full audiometric variable data

| **PHQ-9 Score** | **Mean (Std. Dev)** |
| --- | --- |
| Aggregate Score | 5.0 (4.8) |
| **PHQ-9 Severity Category** | **Count (% of total)** |
| None | 1755 (61.5) |
| Mild | 696 (24.4) |
| Moderate | 237 (8.3) |
| Moderately Severe | 88 (3.1) |
| Severe | 74 (2.8) |

PHQ-9: Patient Health Questionnaire-9.

### Model Performance

On the held-out test sets, the optimized models produced MAE values of 3.03 (CI 95%: 2.91-3.14) and 2.55 (CI 95%: 2.48-2.62) on datasets with audiology-only predictors and all predictors, respectively (**Table 2**).

**Table 2.** LightGBM algorithm cross-validation scores and test set performance

| **Experiment** | **MAE (CI 95%)** |  |
| --- | --- | --- |
|  | **Inner Loop (CV)**<br>**Performance** | **Outer Loop (Test)**<br>**Performance** |
| (1) Audiology Predictors Only | 3.15 (3.10-3.20) | 3.03 (2.91-3.14) |
| (2) All Predictors | 2.57 (2.52-2.61) | 2.55 (2.48-2.62) |

**CV:** Cross-validation; **MAE:** mean absolute error

### Model Interpretation

The top twenty most influential predictors were extracted using the SHAP for both experiments from the first test fold models (**Figure 1** - all predictors; **Figure 2** - audiology predictors only). A thematic analysis of the predictors demonstrated the presence of anxiety, physical limitations, and difficulty concentrating as the top three most influential for predicting PHQ-9 total scores (**Table 3**). The functional audiology predictor *“How often does your hearing cause you to feel frustrated when talking to members of your family or to friends?”* was the fifth-most influential of all variables. A participant’s response of “never” to this predictor was a negative influence on total PHQ-9 total score.

**Figure 1.**
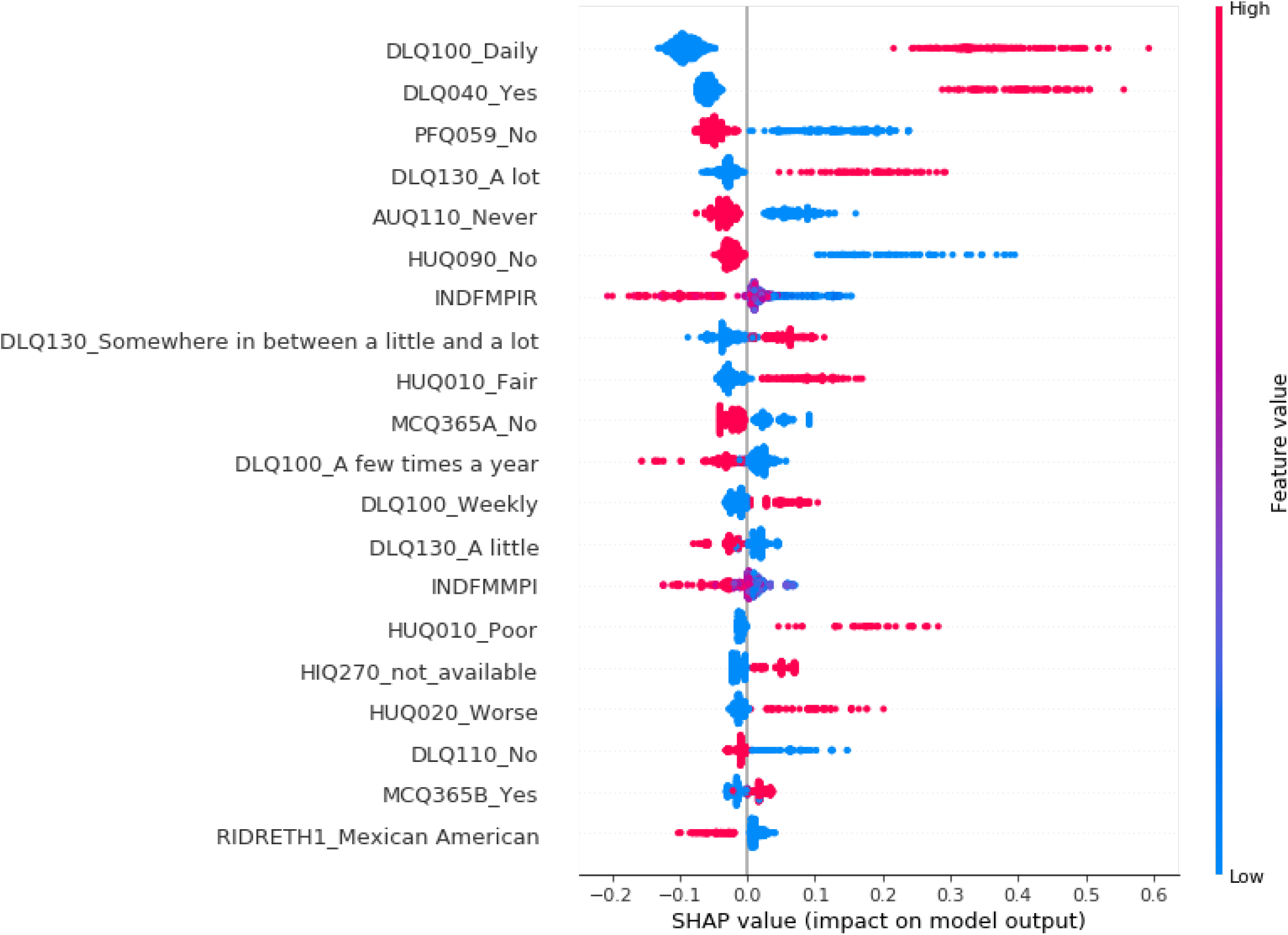
SHapley Additive explanations (SHAP) summary plot depicting the top 20 most influential predictors in predicting PHQ-9 total score from the first test fold model. The x-axis indicates the impact on the target variable. The feature value legend codifies the value of the individual predictor or predictor level. For predictors with “_{text}”, the text indicates the specific level within the categorical predictor. The categorical predictors were one-hot coded, so a ‘high value’ indicates the presence of the labeled level

**Figure 2.**
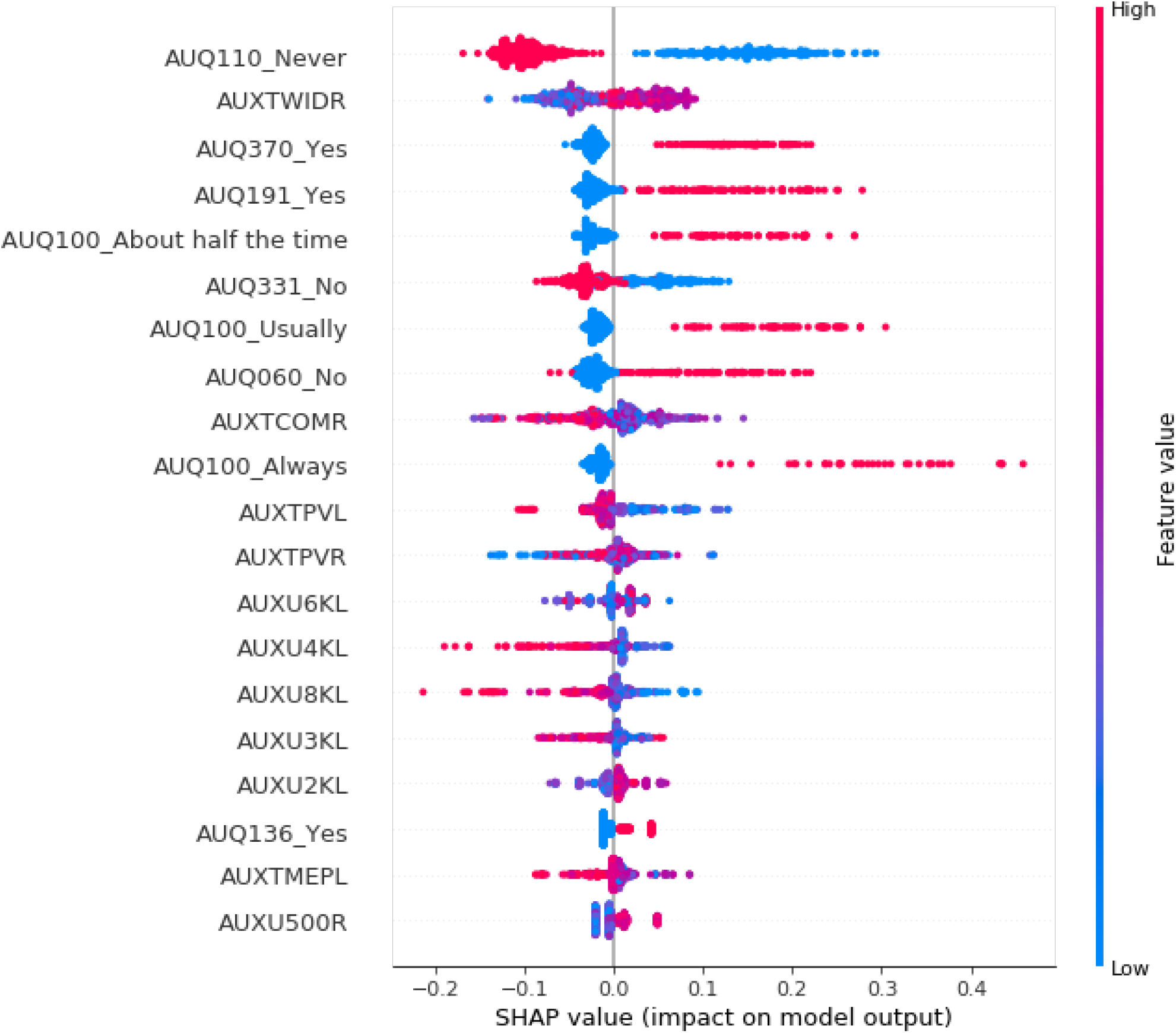
SHapley Additive exPlanations (SHAP) summary plot depicting the top 20 most influential audiometric predictors in predicting PHQ-9 total score from the first test fold model. The x-axis indicates the impact on the target variable. The feature value legend codifies the numeric value of the individual predictor or predictor level. For predictors with “_{text}”, the text indicates the specific level within the categorical predictor. The categorical predictors were one-hot coded, so a ‘high value’ indicates the presence of the labeled level

**Table 3.**
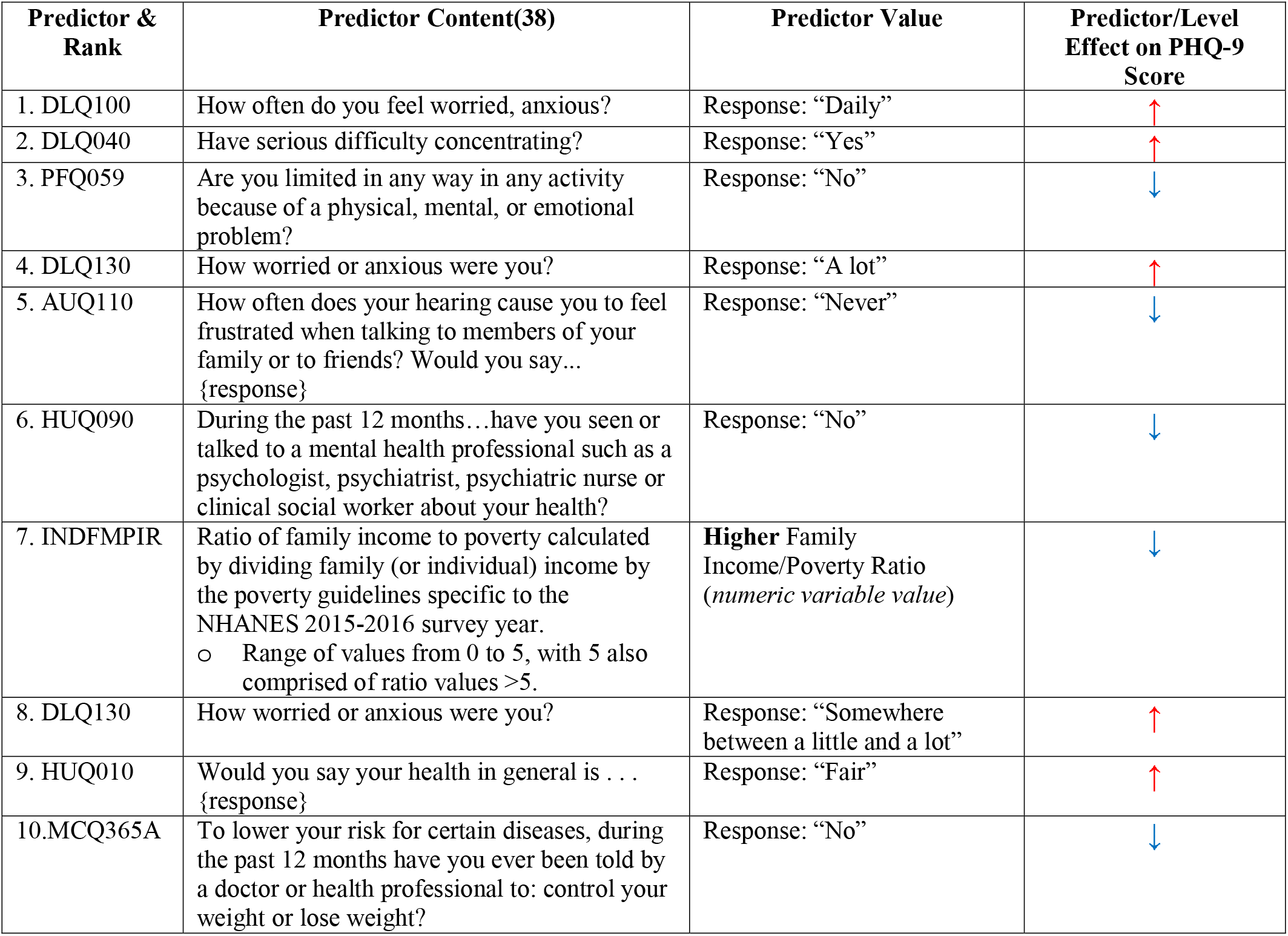
Thematic Analysis of the top ten most influential predictors among all predictors determined by SHapley Additive exPlanations (SHAP); (derived from Figure 1)

In the audiology experiment, the ‘frustration’ variable identified during the all-predictors experiment was the most influential. Following this, predictors pertaining to tympanometric width, exposure to loud sounds, presence of tinnitus, and respondents finding it *“difficult to follow a conversation’”* were the second through fifth most influential, respectively (**Table 4**). Of the top ten most influential audiometric predictors, five were related to social contexts, two for significant noise exposure, two were objective audiometric parameters, and one pertained to presence of bothersome tinnitus.

**Table 4.**
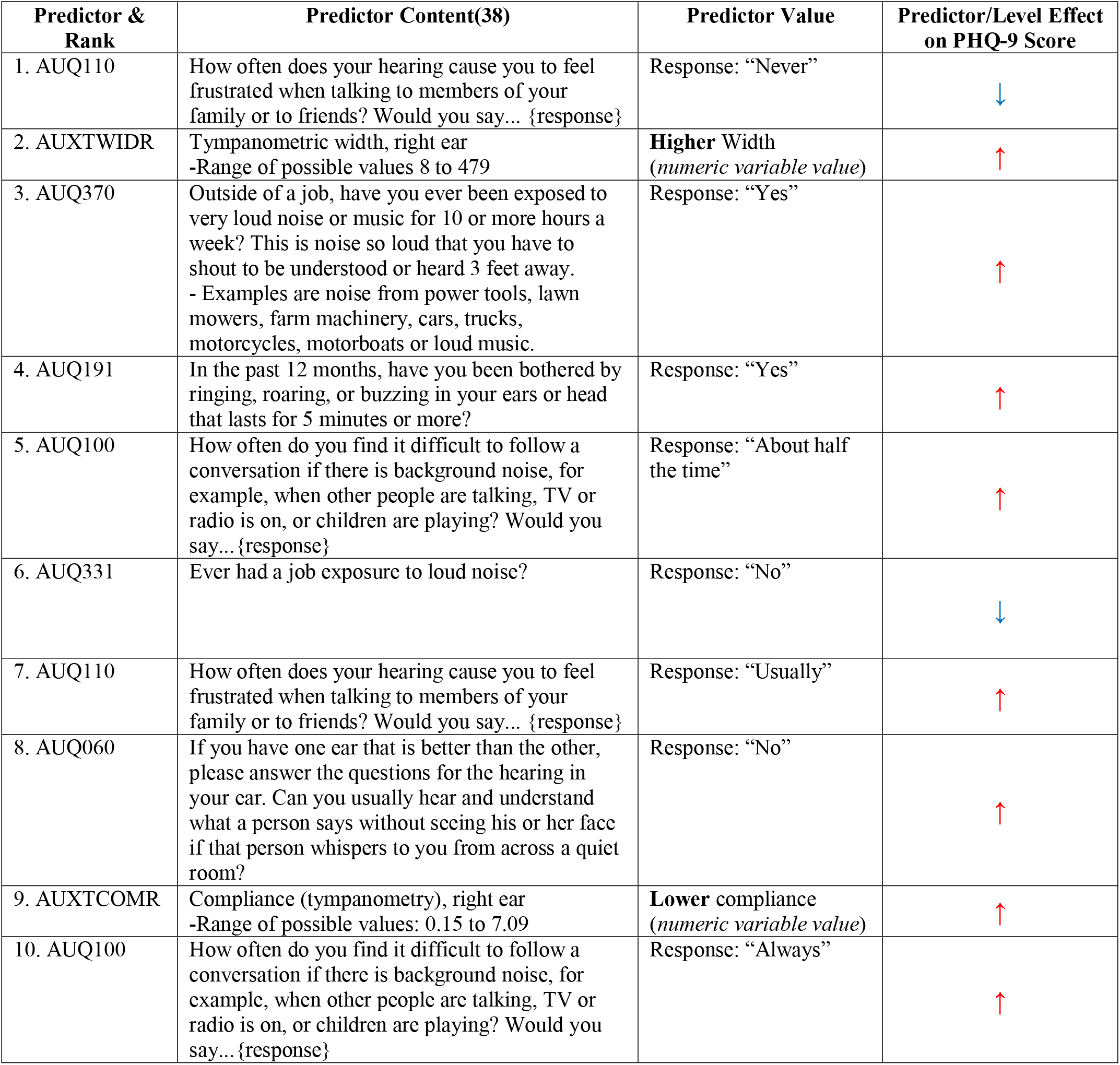
Thematic Analysis of the top ten most influential audiology-only predictors determined by SHapley Additive exPlanations (SHAP) (derived from Figure 2)

## DISCUSSION

In a sample of participants from the 2015-2016 NHANES survey cycle, a supervised machine learning approach successfully identified validated PHQ-9 depression scale scores using audiometric and health determinant predictors. When only audiometric variables were included, there was a small drop in prediction performance on the test set. The most influential audiometric predictors of higher scores on the PHQ-9 depression scale were functional dimensions and not objective audiometric testing. Among the most influential predictors, half were related to social contexts. The remaining were a mixture of content related to noise exposure, tinnitus, and objective audiometric testing. When expanding to include predictors ranging from demographics to other medical and health status content, a social context of hearing loss ranked in the top five most influential variables. These findings highlight relationships related to hearing loss and depression that are understudied and warrant further investigation.

A strong association between social isolation and hearing loss have been demonstrated in older adult populations.(8-10) Greater hearing loss was associated with increased odds of social isolation in women aged 60 to 69 years with an odds ratio of 3.49 (95% confidence interval of 1.91-6.39) per 25-dB of hearing loss in NHANES data from 1999 to 2006.(8) From a functional perspective, subjective hearing assessments correlated more strongly than objective audiometric testing with social isolation in males older than 65 years-old at a U.S. Veterans Administration Medical Center in New York.(10) This result was explained by a belief that subjective assessments capture more aspects of communication including emotional responses and the individuals’ own opinions about negative impact on interpersonal interactions. The authors emphasized that as consequences of hearing loss are recognized by the individual, the sense of isolation, loneliness and inferiority is greater.(10) The observation that hearing loss leads to social isolation is intuitive given that hearing loss leads to impaired efficiency of communication. The connection between social isolation and depression is a natural extension. Our data advances the understanding of this relationship by demonstrating that the social dynamics of hearing loss are highly influential in predicting depression.

If we treat hearing loss, we might reasonably expect prevention of or improvement in social isolation and depression. However, treating hearing loss has not consistently demonstrated improvement in social isolation(26,46), or depression.(25–28) Our study included predictors of self-reported hearing aid or assistive device use, yet none of these predictors were among the most influential. A lack of a clear association may be explained by the inability of hearing aids or devices to improve hearing performance in social settings. At a rudimentary level, hearing devices amplify ambient auditory signals. Basic amplification can and do help individuals with hearing loss. However, sound amplification in context of high levels of excess ambient noise (e.g. at a restaurant or a crowd of simultaneous talkers) may only serve to amplify the cacophony and fail to capture the signal of interest. More advanced modern hearing devices incorporate processing technologies to better isolate the signal from the noise in different social contexts. As of the writing of this manuscript, no data have been published to explore the direct benefit of enhanced auditory signal processing on social isolation or depression. Innovation in hearing devices to produce measurable improvements in social dynamics is important for future investigation and justification of their expense, or study of populations who were known to be using these more advanced technologies.

NHANES data is advantageous for investigations seeking to analyze the interplay between determinants of health and medical conditions. NHANES data has been used to examine the incidence, prevalence and various associative factors with hearing loss.(29–31,31–37) Such factors included associations between hearing loss and hospitalization,(33) poor self-reported health(33), second-hand smoke,(34) falls,(35) occupations with high workplace noise exposures,(36) and lack of hearing protective device use.(36) We used supervised machine learning not just to minimize prediction error, but rather to take advantage of an interpretable non-linear algorithm that handles complex data well. Our tree-based model identified influential predictors that have known associations with depression – self-reported anxiety, difficulty concentrating, and self-perceived limitations due to physical, emotional, or mental ‘problems.’(47–49) By reproducing the influence of these predictors on depression, we are afforded confidence in our findings that the audiometric variables also influence depression. However, this study is not designed as an inferential analysis and further study is necessary to explore these new hypotheses.

Our analytic approach has several limitations that need mentioning. There are well documented limitations of the NHANES dataset that include responder and non-responder bias.(38) We anchored our cohort to participants who had a full audiometric test battery. In doing so, we may have inadvertently introduced selection bias if there was systematic variation in NHANES participants who underwent audiometric testing.

Moreover, this cohort was missing predictor data for the other included predictors (e.g. demographics, medical conditions). We imputed missing numerical data using the predictors’ mean values. Some readers may challenge the omission of certain that may have helped improve predictive performance. It is plausible that other cognitive and/or psychiatric variables may influence the relationship between hearing loss, social relationships and depression. Indeed, there is evidence in cohort analyses linking cognitive decline and hearing loss.(4–7) However, we were not able to explore participants’ cognitive abilities or other psychiatric comorbidities given a lack of valid proxy variables or survey instrument data in the 2015-2016 NHANES dataset. Last, we did not weight our sample to account for the complex survey design of NHANES.(38) The decision was due to, in part, the use of certain predictors that were purely descriptive. The predictors values may not be representative of the nationally representative sample intended by the NHANES study design.

Our analysis demonstrated that the 2015-2016 NHANES dataset is useful for training machine learning models to accurately predict validated PHQ-9 depression scale scores. Such models could be useful in predicting depression scale scores at the point-of-care in conjunction with a standard audiologic assessment. The most influential audiometric predictors of higher scores on the PHQ-9 depression scale were functional dimensions and not objective audiometric testing. Among these influential functional domains, the effect of hearing loss on social relations ranked prominent. Prior research into the effect of hearing aids on depression and social isolation have failed to produce consistent benefits. Our predictive analysis generates a new hypothesis that simply amplifying sound alone, in general, fails to address or improve social dynamics. Recognizing that social aspects of hearing loss may carry more influence in depression than objective audiometry, the efficiency of aural rehabilitation strategies should include outcomes on social dynamics. An inferential analysis is currently in design to explore this hypothesis more rigorously.

## Data Availability

The NHANES database is freely available to the public -- URL: https://wwwn.cdc.gov/nchs/nhanes/

## Manuscript Submission

1. Each of the authors indicated above have contributed to, read and approved this manuscript.
2. **FINANCIAL DISCLOSURE**: no authors have disclosures related to this manuscript.
3. **CONFLICT DISCLOSURE**: no authors have conflicts related to this manuscript.
4. In consideration of the journal reviewing and editing my submission, the authors undersigned transfers, assigns and otherwise conveys all copyright ownership in the event that such work is published.

## ACKNOWLEDGEMENTS

-None-

